# Caregivers’ understanding of sickle cell disease: Knowledge, attitudes, and practices at Masafu general hospital, Uganda. A survey

**DOI:** 10.1101/2025.11.10.25339926

**Authors:** Walter Tevin Kiirya, Jimmy Kizza, Innocent Ayesiga

## Abstract

**Background:** Sickle cell disease (SCD) is a genetic blood disorder that affects approximately 20 million people worldwide, with a relatively high prevalence in sub-Saharan Africa (SSA). Caregivers’ understanding of SCD is crucial to managing this condition. They can be potential stewards of increasing SCD awareness in communities if they are equipped with the correct information. Therefore, the study examined knowledge, attitudes, and practices regarding SCD among caregivers of children receiving chronic care at Masafu General Hospital.

**Methods:** One hundred caregivers were recruited for a cross-sectional survey. The survey used interviewer-administered questionnaires to collect the data. This was then entered into Epidata Manager Version 4.6.0.6 and exported to STATA version 17.0 for analysis.

**Results:** Among 100 sampled people, 80% were female. Most (98%) had heard of SCD, mainly from health facilities (86%). Nearly half (43%) didn’t know that both parents with SCD don’t guarantee a normal child, and 15% of caregivers were unaware of SCD’s cause. A large majority (93%) hadn’t tested their haemoglobin, and 95% didn’t know their partners’ status. Twenty-two per cent believed SCD prevents normal work, perceiving sufferers as weak.

**Conclusion:** The study results indicate the need for continuous caregiver education and increased SCD awareness campaigns in Uganda communities, with strategies to encourage more male involvement.

## Introduction

Sickle cell disease (SCD) is a genetic blood disorder that affects approximately 20 million people worldwide, with a relatively high prevalence in sub-Saharan Africa (SSA), according to the National Heart, Lung, and Blood Institute (NHLBI) 2023[1]. The estimated number of new-borns with SCA globally will increase from 305,800 in 2010 to 400,200 in 2050[2]. More than 75% of the global sickle cell anaemia (SCA) burden occurs in sub-Saharan Africa (SSA). The actual mortality rates are unknown, but approximately 50% to 90% of infants born with SCA in SSA die before their 5th birthday. In Uganda, 20,000 babies per year are thought to be born with SCD, and the high national burden of sickle cell disease (0.7%) was confirmed among a cohort of samples co-tested with HIV [3]. Another study revealed that 1.7% of children in Eastern Uganda tested positive for SCD [4].

Caregivers of children with SCD play a crucial role in managing their condition, including administering medications, recognising and managing symptoms, and accessing appropriate medical care. However, their level of knowledge, attitudes, and practices regarding SCD can have a significant effect on the outcomes of affected children. In addition, they can be potential stewards of increasing SCD awareness in communities if they are equipped with the correct information.

A study conducted in Kinshasa, Congo, revealed that the majority of caregivers felt that society in general had negative perceptions, attitudes, and knowledge about SCD. They reported that children with sickle cell disease are often marginalised, ignored, and excluded from society or school [5]. Another study in the Lubaga division of Kampala, Uganda, reported that 44.2% of the participants did not know the cause of SCD [6]. Another study revealed that 8% of household respondents in Eastern Uganda believed that it was a curse/punishment from God compared with 2% in the West [7].

Masafu General Hospital (MGH) is the only public general hospital in Busia district, Eastern Uganda, that provides chronic care services to approximately 400 children with SCD, as stipulated by the Ministry of Health guidelines, including monthly review visits and malaria and anaemia prophylaxis, among others. Previous studies have shown a lack of knowledge and negative perceptions of sickle cell disease among the general public. However, there is likely a critical knowledge gap regarding caregivers’ knowledge, attitudes, and practices regarding sickle cell disease among children receiving chronic care at Masafu General Hospital, Busia district, Uganda. Existing research has focused primarily on other countries and often lacks a localised perspective.

Therefore, this survey aimed to determine caregivers’ knowledge, attitudes, and practices regarding SCD among children receiving chronic care at Masafu General Hospital. The evidence generated from this research will inform policy development and program implementation to improve the care and management of SCD in Uganda and other settings. By understanding caregivers’ knowledge, attitudes, and practices, healthcare policies and programs can be tailored to address specific challenges, promote culturally sensitive care, and enhance access to quality healthcare services for children with SCD, thereby improving health outcomes and reducing the burden of the disease.

## Materials and methods

A descriptive cross-sectional survey was used to assess the knowledge, attitudes, and practices among caregivers of children with SCD receiving chronic care. The survey was conducted at Masafu General Hospital, a general public hospital in the Eastern Region of Uganda, Busia district, located approximately 195 kilometres from Kampala, the capital city, from 1^st^ January 2024 to 31^st^ August 2024 as the overall research period, including planning, preparations, data collection, and other activities. Caregivers of children with SCD attending chronic care at Masafu General Hospital SCD clinic. About 15-20 caregivers of children attend the routine Tuesday clinic weekly. Convenience sampling was used, and study participants were enrolled as they arrived at the facility for routine visits and reviews. We included both males and females, as well as guardians/parents aged 18 and above, who were willing to provide written informed consent. We excluded those younger than 18 years. Eligible participants were interviewed during routine clinic visits every Tuesday from May 1st to June 30th, 2024. A research assistant and researcher administered the questionnaire after providing informed consent. The questionnaire was pretested to ensure it aligned with the study objectives. The dependent variables in this study included caregivers’ knowledge, attitudes, and practices related to sickle cell disease. Questions about the source of information on SCD, its cause, symptoms, conditions that worsen the disease/triggers, and routine drugs prescribed to affected children by clinicians were used to assess participants’ knowledge of SCD. Practices of premarital screening or knowledge of haemoglobin phenotype status, and perceptions of people with SCD in the community, were also evaluated. The independent variables included education level, occupation, gender, marital status, and access to information. A sample size of 100 caregivers was determined using Kish’s formula (Kish, 1965). With the expected prevalence of knowledge of SCD and positive attitudes among caregivers at 7% in a study in Congo,^5^ the desired precision is 5%, and a confidence level of 95% was used. An estimated sample size of 100 participants was obtained for this survey. Data were collected via a data collection tool (questionnaire) that was administered by trained research assistants at the Sickle Cell Clinic, Masafu General Hospital. This was then entered into Epidata Manager Version 4.6.0.6 and exported to STATA version 17.0 for analysis. We report the means and standard deviations (SDs) for continuous variables and proportions for categorical variables. Some of the data is visualised via bar graphs.

## Results

### Description of the study participants

We recruited 100 participants: 80 females and 20 males. They were mainly the children’s mothers and fathers. Most participants (43%) were aged 36-45, followed by those aged 18-35 (40%). 90% of the participants were married; the majority (44%) had attained primary education as their highest level of education, and farming (52%) was the occupation of most of them. The mean number of children among participants was 5.11 (SD = 2.68), and the mean number of children with SCD was 1.34 (SD = 0.75). (Table 1).

**Table 1:** Key demographic characteristics of the study participants (n=100)

| Parameter |  | Percentage (%) |
| --- | --- | --- |
| **Age (years)** | 18-35 | 40 |
|  | 36-45 | 43 |
|  | 46-60 | 12 |
|  | Above 60 | 5 |
| **Gender** | Male | 20 |
|  | Female | 80 |
| **Marital status** | Married | 90 |
|  | Not married | 10 |
| **Education level** | No education | 13 |
|  | Primary | 44 |
|  | Secondary | 30 |
|  | Tertiary | 13 |
| **Occupation** | Health | 2 |
|  | Farming | 52 |
|  | Teaching | 8 |
|  | Business | 30 |
|  | Other | 8 |

### Knowledge of caregivers about SCD

98% of the participants had heard of sickle cell disease, with the majority (86%) reporting receiving this information from health facilities/centres. (Fig. 1)

**Fig. 1:**
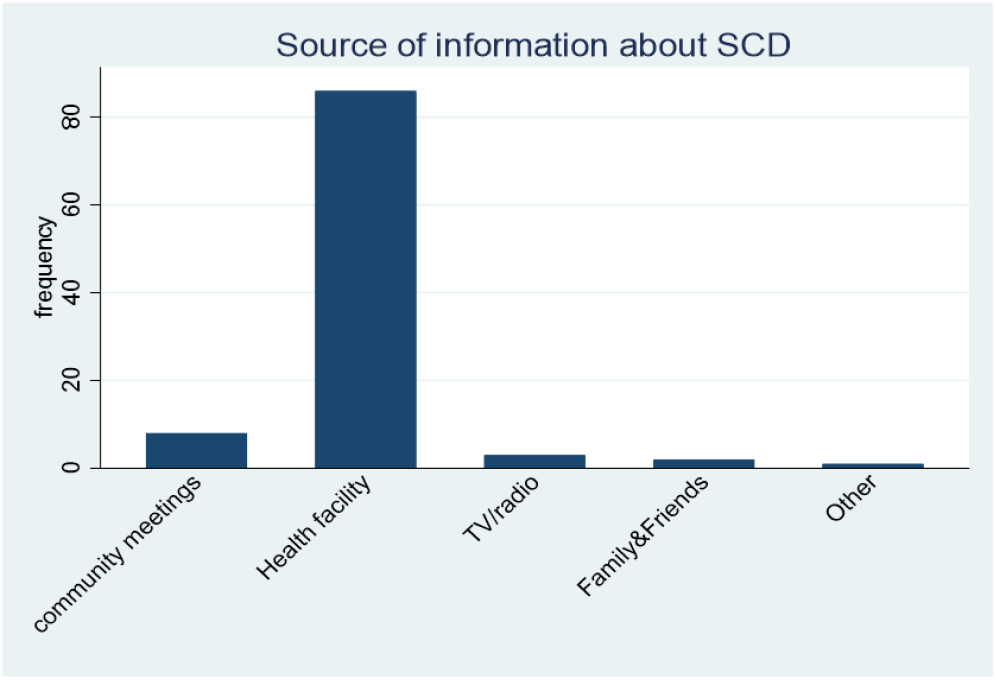
Bar graph showing the source of information about SCD.

Health facilities are the major source of information about sickle cell disease (SCD) among caregivers of children with SCD receiving chronic care. 85% of the participants reported that SCD is a familial or inherited disorder, with 95% reporting that SCD is diagnosed through a “blood test”. In addition, the vast majority (86%) of the participants reported that there was no cure for SCD; however, 14% reported that there was a cure for the condition, referring to traditional medicine. Yellow eyes, paleness of the body, and pain in the limbs were the most common symptoms of SCD among children. In contrast, Fansidar, folic acid, and hydroxyurea were the most common drugs routinely given to children with SCD by caregivers as prescribed by the clinicians. Participants also noted that coldness, cough, and fever were the most common triggers of SCD crises in children. (Table 2)

**Table 2:** Table showing the symptoms of SCD, triggers for crises, and routine drugs for management listed by the study participants.

| Parameter |  | Frequency(N=100) |
| --- | --- | --- |
| Symptoms of SCD | Yellow Eyes | 85 |
|  | Paleness of the body | 72 |
|  | Pain in the limbs | 50 |
|  | Frequent sickness | 28 |
| Triggers for SCD crisis | Coldness | 84 |
|  | Fevers | 10 |
|  | Cough | 41 |
|  | Strenuous activity | 7 |
|  | Dehydration | 2 |
| Routine drugs for SCD | Fansidar (SP) | 62 |
|  | Folic acid | 93 |
|  | Hydroxyurea | 25 |
|  | Pen V | 8 |
|  | chloroquine | 12 |
|  | Analgesics | 9 |

More than half of the participants (57%) mentioned that there was no chance of producing a normal child if both parents had SCD, 3(3%) mentioned “all children would be normal”, 7(7%) mentioned a half chance that the child would be normal, and 30 (30%) participants did not know. (Table 3)

**Table 3:** shows the responses of the study participants to the question of the chances of producing a normal child when both parents have SCD.

| If both parents are sick with SCD, what are the chances of producing a normal child? | Frequency | Percentage |
| --- | --- | --- |
| None of the children | 57 | 57.00 |
| All the children | 3 | 3.00 |
| Half a chance for the children | 7 | 7.00 |
| Quarter chance the child will be normal | 3 | 3.00 |
| Don't know | 30 | 30.00 |
| **Total** | 100 | 100.00 |

### Practice and attitudes towards Sickle Cell Disease

Most (93%) of the caregivers of children with SCD had not tested for their haemoglobin phenotype, and almost all participants (95%) did not know their partner’s haemoglobin phenotype. The majority (79%) stated that the reason for testing, if they were to do so, was “to know the status”; 16 (16%) of the participants said “because the child has SCD”, and 3 did not wish to test for SCD (Fig. 2).

The primary reason given by caregivers of children, if they were to test, is to know their sickle cell disease (SCD) status, and a significant number of them because their children had this condition. 22% of the participants reported that a person with SCD could not perform/work as usual as other individuals because they were perceived as weak or lacking energy.

**Fig. 2:**
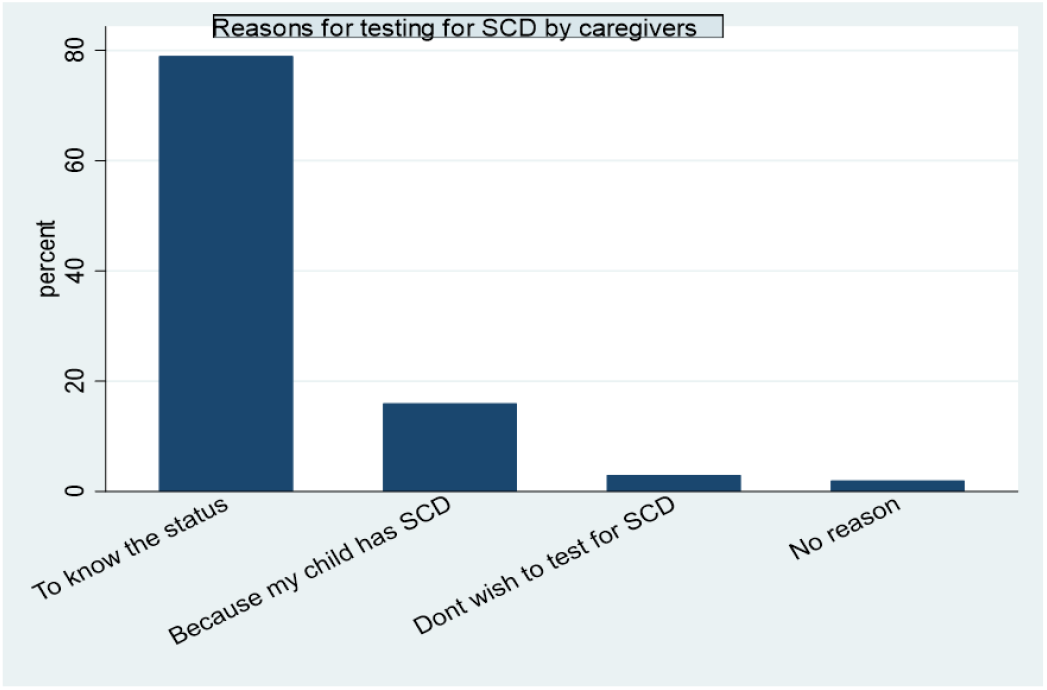
Bar graph showing the reasons for testing for SCD by caregivers.

## Discussion

Most studies conducted in Uganda have placed greater emphasis on the community at large, with less focus on caregivers in particular, probably because they are always attending SCD clinics with their children, among other reasons. However, they play a vital role in managing the condition and can be potential stewards to increase SCD awareness in communities if equipped with the correct information. Overall, the findings revealed that the majority of caregivers of children with SCD at Masafu General Hospital had poor knowledge, practices and perceptions/attitudes, such as failure to test for haemoglobin phenotype status by most (93%) of the participants, indicating gaps in knowledge, a more profound understanding of the condition, and clinical practice.

This survey targeted adult caregivers, both male and female. This campaign attracted a lower proportion of males. This may be because most men were away from work, as highlighted by Francis et al. 2008[8]. However, this may also be because of the community perception that it’s the mother’s cultural role to take children to the hospital, exempting fathers, and insufficient outreach efforts to men in society. The study results are similar to those of a study that also highlighted that more women utilise health facilities/services in Wakiso District [9]. This may continue to underscore community campaigns, since men are often the decision-makers in most Ugandan homes. Some initiatives to increase male involvement may include organising male-targeted education sessions outside regular hospital or work hours, such as weekends and evenings; engaging male community leaders and role models, such as religious figures; and partnering with employers to deliver brief SCD education and screening programs at workplaces.

Most participants had heard of SCD, suggesting they were aware of its existence. The majority, having obtained information from health facilities, may indicate increased effort in health care settings/systems to inform the public of SCD. However, the small proportion who received such information from community meetings may suggest limited community engagement efforts regarding SCD in this setting. These results differ from those reported by a study, which highlighted that a small proportion of community participants had received SCD from health facilities [6]. Another study reported that schools ranked highest in Ghana for sources of knowledge about sickle cell disease, whereas health centres ranked highest in our study [10]. This evidence highlights the need to promote greater health education about sickle cell disease in communities and families.

In addition to having heard of the disease, a majority knew the causes, signs, symptoms, and triggers of SCD crises, which may aid prompt care-seeking, as caregivers already see the warning signs. Yellow eyes, paleness of the body, and pain in the limbs were the most common symptoms of SCD among children. In contrast, Fansidar, folic acid, and hydroxyurea were the most common drugs routinely given to children with SCD by caregivers as prescribed by the clinicians. Participants also noted that coldness, cough, and fever were the most common triggers of SCD crises in children. The results could also inform which information should be included in SCD awareness campaigns. Almost half of the participants did not know that there was no chance of producing a normal child if both parents had SCD. This demonstrated a knowledge gap in the basics of the condition’s pathophysiology, which can reduce efforts to achieve early diagnosis and informed family planning decisions, increasing the burden of the disease in the population.

Out of 100 caregivers, only seven were aware of their haemoglobin phenotype status; the majority stated that the reason for testing, if they were to do so, was “to know the status”; some said” because the child has SCD” (16%), and a few did not wish to test for their haemoglobin phenotype status. In addition, the majority of the caregivers did not know their partner’s sickle cell disease status. They had a poor attitude towards preventing the propagation of SCD, and this should serve as a spot for a very aggressive campaign on the importance of premarital screening. 22% of the participants reported that a person with SCD could not perform/work as usual as other individuals because they were perceived as weak. This may indicate possible stigmatisation/discrimination of people with SCD in the community, which may significantly affect their mental health. This finding is similar to that of other researchers, who reported that individuals with SCD often report SCD-related stigma [11-12].

## Conclusions

Overall, the majority of caregivers of children with SCD at Masafu General Hospital had generally poor knowledge about SCD, with notably fair knowledge about the symptoms and triggers of SCD crises. More than half of the participants had poor practices regarding SCD, with most (93%) unaware of their own haemoglobin phenotype and 95% unaware of their partners’ haemoglobin phenotype. There was poor perception towards SCD, where 22% of our participants reported that a person with SCD cannot perform/work as usual, like other individuals, because they were perceived as weak.

### Recommendations

Given that the caregivers of children with SCD reported poor practices towards SCD, we recommend continuous medical education sessions to all caregivers on SCD, pathophysiology, screening, and other related information. To reinforce knowledge and serve as quick reminders, using health information tools, such as posters, can be highly effective. Placing these posters on hospital notice boards can provide caregivers with readily accessible information regarding SCD and screening, even in the local language. Further research is needed in this area too. The Ministry of Health and other stakeholders should strengthen mass sensitisation programs in communities about SCD, highlighting its causes, symptoms, and the importance of screening to improve people’s practices and prevent possible stigma/discrimination against those affected. There is a need to formulate strategies to encourage male involvement in SCD campaigns.

## Data Availability

All data produced in the present study are available upon reasonable request to the authors

## Declarations

## Data Availability Statement

Data from the datasets analysed in this study are available from the authors upon reasonable request.

## List of Abbreviations

MGH: Masafu General Hospital
NHLBI: National Heart, Lung, and Blood Institute
SCA: Sickle Cell Anaemia
SCD: Sickle Cell Disease
SD: Standard Deviation
SP: Fansidar
SSA: Sub-Saharan Africa

## Author Contributions

WTK conceptualised the study WTK coordinated data collection

WTK and JK coordinated data analysis WTK developed the initial manuscript draft

IA provided technical input to the manuscript

All authors reviewed the initial manuscript and approved the relevant changes before submission for publication.

## Clinical Trial Number

Not Applicable

## Acknowledgements

We acknowledge the administration of Masafu General Hospital for the necessary support provided, and we extend special thanks to the medical team of the SCD clinic at MGH.

## Ethical Considerations

The School of Biomedical Sciences Research and Ethics Committee of Makerere University approved the study procedures under grant number SBS-2024-524. Written informed consent was obtained from all participants before enrolment into the study. The survey adhered to the Declaration of Helsinki in all its activities.

